# Antimicrobial resistance genomics across Africa: critical determinants, repository bias and regional coordination

**DOI:** 10.64898/2026.08.31.26361859

**Authors:** Ruth Omani, Grace N. Maina, Folorunso Oludayo Fasina

## Abstract

Public genomic repositories can support antimicrobial resistance (AMR) surveillance, but unequal sampling can bias interpretation. We characterised AMR determinants, multicountry genomic cluster overlap and surveillance gaps across Africa using an NCBI Pathogen Detection snapshot retrieved on 24 August 2026 for 55 African Union member states. Records were validated and deduplicated by BioSample, and complete AMRFinderPlus calls were summarised across five United Nations M49 subregions and eight overlapping regional economic communities (RECs). Country-pair cluster overlap was assessed using the Jaccard index, while project-based and composition-standardised sensitivity analyses evaluated repository bias. The dataset contained 86,829 unique BioSamples from 51 states; South Africa, Malawi and Kenya contributed 55.8%. Complete extended-spectrum β-lactamase calls were detected in 21,513 isolates and carbapenemase calls in 4,642. *bla*CTX-M-15 dominated the ESBL profile, while NDM and OXA types predominated. Seventy clusters contained carbapenemase-positive isolates from at least two countries. A shared REC covered all participating countries in 38 clusters, while 32 crossed REC boundaries. Normalised country-pair overlap was low, with a maximum Jaccard index of 9.5%. Project balancing reduced the Northern African carbapenemase estimate from 32.3% to 17.9% and the Eastern African ESBL estimate from 36.9% to 12.5%. Public repositories identify determinants and clusters for investigation but do not estimate prevalence or transmission. AMR surveillance should combine national confirmation, regional institution-led investigation where countries share an REC, and continent-wide coordination through Africa CDC for cross-REC signals, supported by representative One Health sampling, standardised metadata and sustained African sequencing capacity.

## 1 Introduction

Antimicrobial resistance (AMR) reduces the effectiveness of essential medicines and threatens health systems, food security and sustainable development. In 2021, bacterial AMR was associated with an estimated 4.71 million deaths globally, including 1.14 million deaths directly attributable to resistance, while approximately 39.1 million attributable deaths are projected between 2025 and 2050 [1]. Africa carries a substantial burden, but uneven laboratory capacity and limited microbiological testing and surveillance constrain understanding of the organisms and resistance mechanisms circulating across the continent [2–4]. This limitation is particularly important because movements of people, livestock and wildlife, together with interconnected food systems, trade routes, transport networks and water systems, connect African countries and create pathways for resistant organisms and mobile resistance determinants to cross national borders. The African Union (AU), comprising 55 member states, recognises eight regional economic communities (RECs) as pillars of regional integration and coordination, providing established structures through which cross-border resistance threats can be addressed [5,6].

Whole-genome sequencing (WGS) determines the nucleotide sequence of most or all of a bacterial genome. In bacteriology, it can identify acquired resistance genes, resistance-associated point mutations and genomic relatedness among bacterial isolates. When combined with phenotypic susceptibility testing and reliable contextual metadata, it can support investigation of AMR across human, animal and environmental sources [7–9]. The National Center for Biotechnology Information (NCBI) maintains public sequence and genome resources in which genome assemblies and associated reference information are collected and curated for reuse [10]. Within this infrastructure, the NCBI Pathogen Detection system supports such analyses by linking assembled genomes to BioSample metadata, identifying resistance determinants using AMRFinderPlus and grouping related isolates into single-nucleotide polymorphism clusters [11,12]. However, repository records originate from heterogeneous research projects and surveillance programmes, and their analysis must account for differences in sampling, organism composition, sequencing activity and metadata completeness. These concerns are especially relevant in Africa, where genomic surveillance has expanded but sequencing and bioinformatics capacity, trained personnel, sustainable funding and genomic data production remain uneven and concentrated in a limited number of countries and institutions [13–17].

This study analysed NCBI Pathogen Detection records from all 55 AU member states to characterise complete AMRFinderPlus calls for extended-spectrum β-lactamases, carbapenemases, *mcr* genes and *mecA* across countries, organisms and One Health sources. Geographic comparisons were conducted using the five mutually exclusive United Nations M49 subregions—Eastern, Middle, Northern, Southern and Western Africa—while the eight AU- recognised RECs were examined as a governance and policy framework because their memberships overlap [6,18,19]. The analysis also examined country-level SNP-cluster connectivity, the influence of BioProject and sample composition on subregional patterns, the relationship between multicountry genomic clusters and REC coordination structures, and inequalities in repository representation and metadata completeness.

## 2 Methods

### 2.1 Study design, data selection and classification

A cross-sectional repository analysis was conducted using an NCBI Pathogen Detection snapshot retrieved on 24 August 2026. The platform links assembled bacterial genomes with BioSample metadata, AMRFinderPlus resistance predictions and SNP-cluster assignments [11,12]. Records were selected for all 55 African Union (AU) member states using geographic-location metadata. Western Sahara was used as the repository label for the Sahrawi Arab Democratic Republic, while Sudan and South Sudan were treated separately. Records required a valid BioSample accession and were deduplicated to retain the most recent and complete record per BioSample. Detailed selection rules and exclusion audits are provided in S4 and S5 Files. Countries were grouped into the five UN M49 African subregions and assigned to the eight AU-recognised regional economic communities (RECs) using memberships current on 24 August 2026 [5,6,19–22]. Overlapping REC membership was retained, and REC totals were analysed separately. Burkina Faso, Mali and Niger were excluded from ECOWAS following their withdrawal in January 2025 [23].

Available collection, host, source, location, BioProject and institutional metadata were retained. Isolates were classified as human/clinical, animal, food or animal product, environmental or unclassified. Primary AMR outcomes were restricted to AMRFinderPlus calls classified as COMPLETE; partial calls and resistance-associated point mutations were retained separately [11,12]. ESBLs included the *bla*CTX-M, *bla*PER and *bla*VEB families and selected SHV variants. Carbapenemases included the *bla*NDM, *bla*KPC, *bla*VIM, *bla*IMP and *bla*IMI families and selected OXA carbapenemases. Complete *mcr* genes and *mecA* were analysed separately. Dual positivity required at least one complete ESBL and one complete carbapenemase in the same BioSample.

### 2.2 Statistical and genomic-connectivity analyses

Counts and repository frequencies were summarised by country, subregion, REC, organism and One Health source. Organism–country percentages were presented only for strata containing at least 50 isolates; smaller strata were reported using exact counts. BioProject concentration was assessed using the largest-project shares, the Herfindahl–Hirschman index and its inverse. Leave-one-project-out analyses examined the influence of individual projects.

Sensitivity analyses used L2-regularised logistic models containing subregion, organism group, source and collection period. Composition-standardised estimates used a common covariate distribution, while project-balanced estimates gave each BioProject equal weight. These estimates assessed the influence of sample and project composition and were not interpreted as population-level prevalence. Geographic differences were not tested inferentially because repository isolates were not probability samples.

A multicountry cluster was defined as an NCBI SNP-cluster root containing isolates from at least two African countries. Country-pair connectivity was measured using the Jaccard index. A strict carbapenemase connection required both countries to contribute a carbapenemase-positive isolate to the same cluster; the broader definition included clusters containing a complete carbapenemase in any isolate. Cluster membership indicated genomic relatedness, not direct transmission or outbreak membership [12].

### 2.3 Policy mapping, reproducibility and ethics

Multicountry clusters were mapped against REC membership. Official AU and REC sources were reviewed for documented AMR frameworks, laboratory networks, One Health programmes and regional health-coordination mechanisms available on 24 August 2026 [6,18,20–22]. The resulting matrix identified institutional mechanisms but did not assess their implementation or effectiveness. Trade-corridor transmission was not examined because mobility data were unavailable.

Data processing and analysis were conducted using reproducible Python pipelines. S1–S5 Files comprise the raw repository snapshot, the cleaned and deduplicated analysis-ready isolate dataset, analytical result tables and policy-source matrix, reproducibility code, and the geographic-validation, BioSample- deduplication and provenance audit. Repository creation dates were treated as deposition dates rather than dates of AMR emergence. The study used publicly accessible, non-identifiable records and involved no participant recruitment or access to individual clinical information.

## 3 Results

### 3.1 Dataset composition and geographic representation

Of 108,729 downloaded records, one record with the location value “Pacific Ocean: Guinea” was excluded because it did not meet the prespecified country-prefix rule. A further 21,899 records sharing a BioSample accession with another record were removed under the one-record-per-BioSample deduplication rule, leaving 86,829 unique BioSamples. Qualifying records were available for 51 of the 55 AU member states; Equatorial Guinea, Lesotho, Seychelles and Western Sahara had no qualifying records. Repository representation was highly uneven: South Africa, Malawi and Kenya accounted for 55.8% of isolates, while 15 countries contributed fewer than 50. Eastern and Southern Africa together comprised 73.5% of the dataset (Table 1; Fig 1).

**Fig 1.**
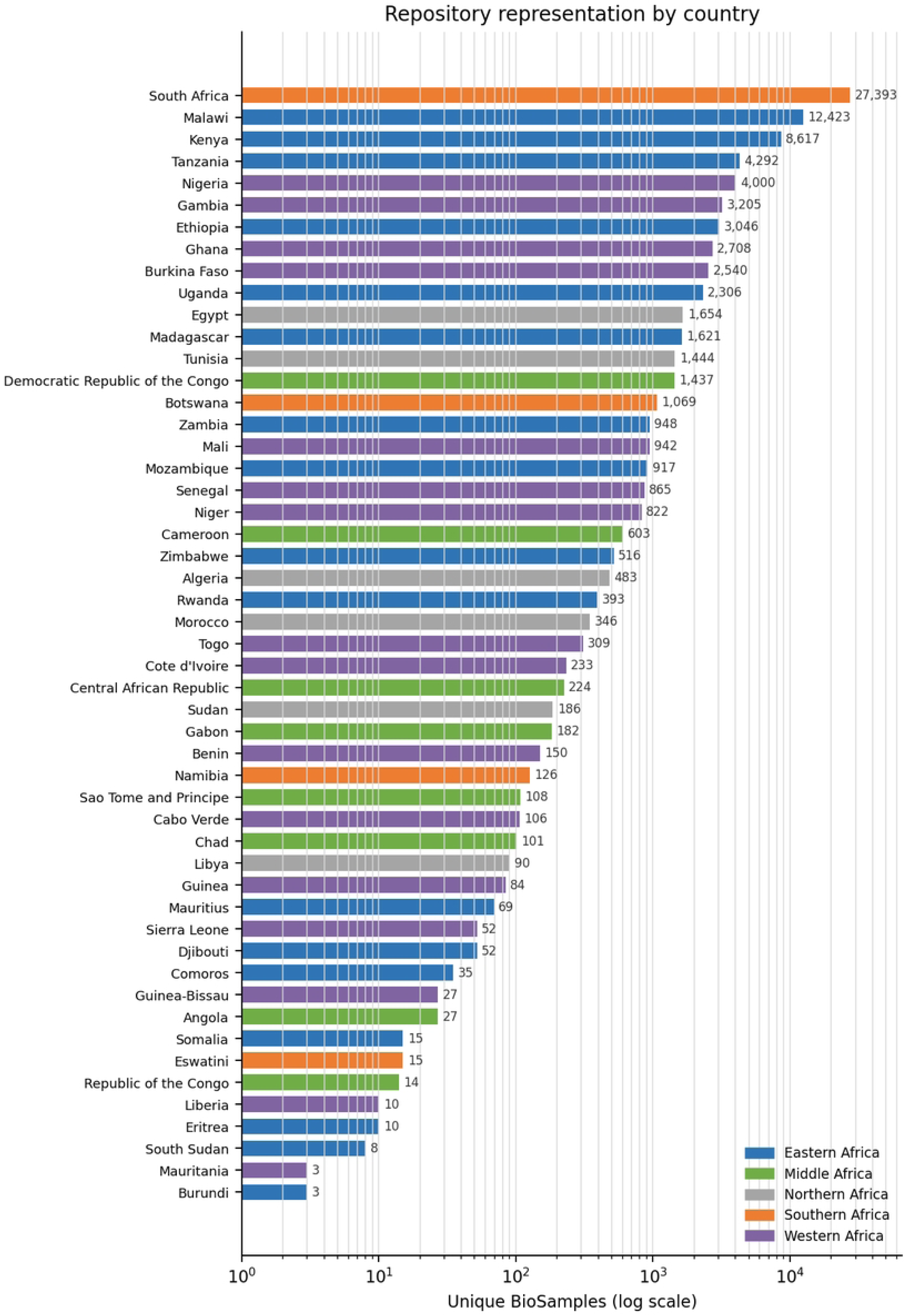
Repository representation among the 51 African Union member states with qualifying records.

**Table 1.** Repository representation and complete priority AMR outcomes by UN M49 subregion.

| UN M49 subregion | Isolates | Africa share | Countries | ESBL + | Carbapenemase + | Dual + |
| --- | --- | --- | --- | --- | --- | --- |
| Eastern Africa | 35,271 | 40.6% | 17 | 13,032 (37.0%) | 646 (1.8%) | 382 (1.1%) |
| Southern Africa | 28,603 | 32.9% | 4 | 3,734 (13.1%) | 1,671 (5.8%) | 1,297 (4.5%) |
| Western Africa | 16,056 | 18.5% | 16 | 2,678 (16.7%) | 966 (6.0%) | 586 (3.7%) |
| Northern Africa | 4,203 | 4.8% | 6 | 1,541 (36.7%) | 1,356 (32.3%) | 756 (18.0%) |
| Middle Africa | 2,696 | 3.1% | 8 | 528 (19.6%) | 3 (0.1%) | 3 (0.1%) |

*Note: Frequencies describe the deposited genomic collection and are not population-level prevalence estimates.*

### 3.2 Distribution of critical AMR determinants

Complete ESBLs were detected in 21,513 isolates (24.8%) and carbapenemases in 4,642 (5.3%). Dual positivity occurred in 3,024 isolates (3.5%), representing 65.1% of carbapenemase-positive isolates. *bla*CTX-M-15 accounted for 80.5% of ESBL-positive isolates, whereas carbapenemase detections were distributed mainly across the NDM and OXA families. Complete *mcr* genes and *mecA* were detected in 262 and 935 isolates, respectively (Table 2; Fig 2).

**Fig 2.**
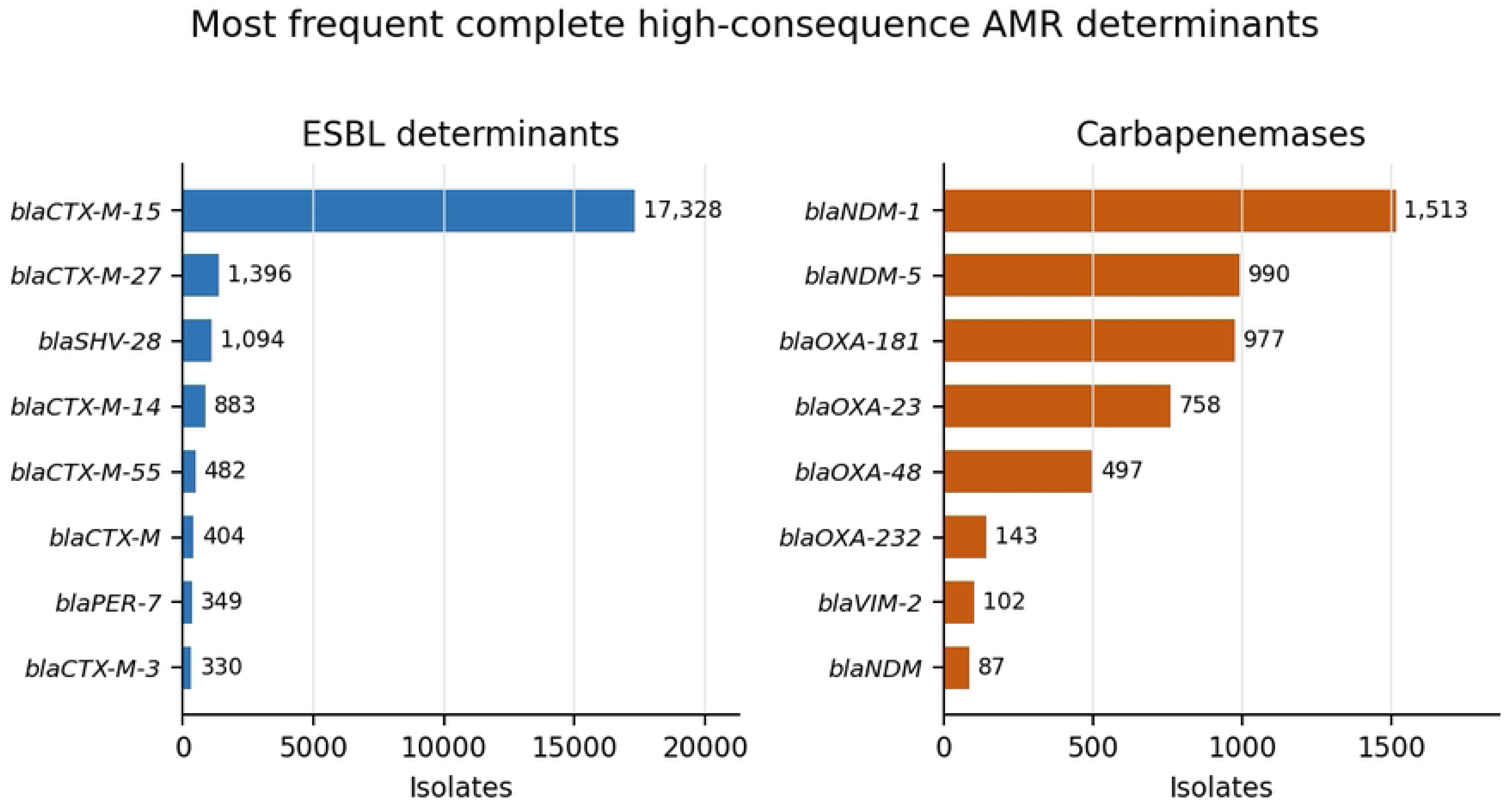
Most frequent complete ESBL genes and carbapenemases in the African repository.

**Table 2.** Most frequent complete high-consequence AMR determinants.

| Class | Determinant | Isolates | Africa repository frequency |
| --- | --- | --- | --- |
| ESBL | *bla*CTX-M-15 | 17,328 | 19.956% |
| ESBL | *bla*CTX-M-27 | 1,396 | 1.608% |
| ESBL | *bla*SHV-28 | 1,094 | 1.260% |
| ESBL | *bla*CTX-M-14 | 883 | 1.017% |
| ESBL | *bla*CTX-M-55 | 482 | 0.555% |
| Carbapenemase | *bla*NDM-1 | 1,513 | 1.743% |
| Carbapenemase | *bla*NDM-5 | 990 | 1.140% |
| Carbapenemase | *bla*OXA-181 | 977 | 1.125% |
| Carbapenemase | *blaOXA-23* | 758 | 0.873% |
| Carbapenemase | *blaOXA-48* | 497 | 0.572% |
| Carbapenemase | *blaOXA-232* | 143 | 0.165% |
| mcr | *mcr-1.1* | 116 | 0.134% |
| mcr | *mcr-10.1* | 66 | 0.076% |
| mcr | *mcr-9.1* | 48 | 0.055% |
| mecA | *mecA* | 935 | 1.077% |

Note: Primary outcomes used COMPLETE AMRFinderPlus calls. Percentages use all 86,829 BioSamples as the denominator; gene counts are not mutually exclusive.

### 3.3 Organism and One Health source patterns

Determinant profiles differed markedly by organism. *Klebsiella pneumoniae* had the highest ESBL repository frequency among the major Enterobacterales (80.6%), and 19.7% carried both an ESBL and a carbapenemase. Carbapenemases were most frequent in *Acinetobacter baumannii* (73.9%) and *Pseudomonas aeruginosa* (30.9%). Almost all *mecA* detections were in *Staphylococcus aureus* (923/935; 98.7%) (Table 3; Fig 3).

**Fig 3.**
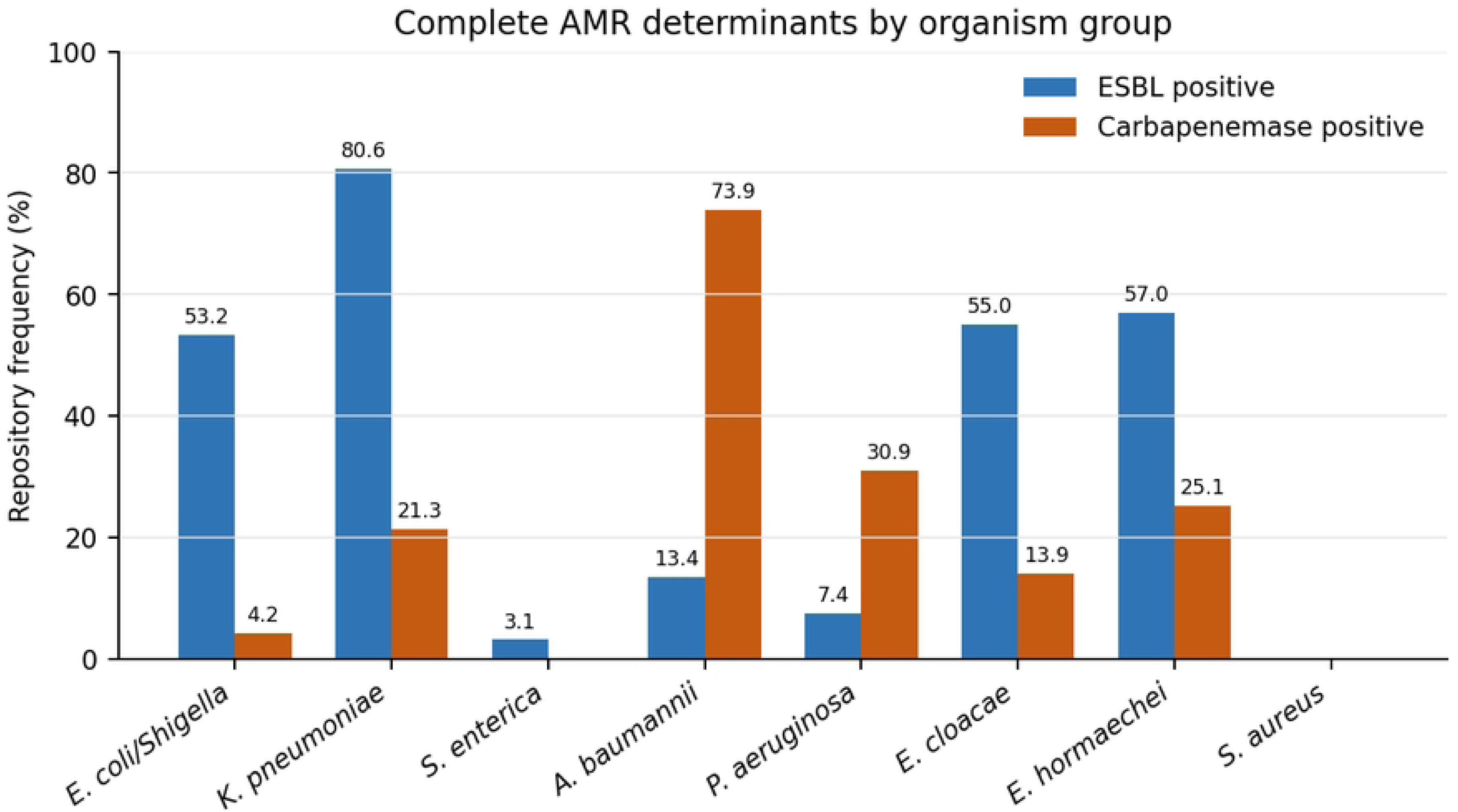
Organism-specific repository frequencies of complete ESBL and carbapenemase determinants.

**Table 3.** Complete priority AMR outcomes in selected organism groups.

| Organism group | Isolates | ESBL + | Carbapenemase + | Dual + | mcr + | mecA + |
| --- | --- | --- | --- | --- | --- | --- |
| *E. coli* and *Shigella* | 21,853 | 11,635 (53.2%) | 917 (4.2%) | 652 (3.0%) | 116 | 1 |
| *Klebsiella pneumoniae* | 10,165 | 8,192 (80.6%) | 2,165 (21.3%) | 1,997 (19.7%) | 20 | 0 |
| *Salmonella enterica* | 18,089 | 560 (3.1%) | 34 (0.2%) | 14 (0.1%) | 12 | 0 |
| *Acinetobacter baumannii* | 1,236 | 165 (13.4%) | 913 (73.9%) | 158 (12.8%) | 2 | 0 |
| *Pseudomonas aeruginosa* | 1,023 | 76 (7.4%) | 316 (30.9%) | 49 (4.8%) | 0 | 0 |
| *Enterobacter cloacae* | 309 | 170 (55.0%) | 43 (13.9%) | 26 (8.4%) | 33 | 0 |
| *Enterobacter hormaechei* | 414 | 236 (57.0%) | 104 (25.1%) | 61 (14.7%) | 22 | 0 |
| *Staphylococcus aureus* | 3,080 | 0 (0.0%) | 0 (0.0%) | 0 (0.0%) | 0 | 923 |

Note: The table is organism-stratified to reduce distortion from differences in pathogen composition between country collections.

Human or clinical isolates comprised 75.6% of the repository, whereas animal, food and environmental isolates together comprised 11.1%. Environmental samples accounted for 293 of the 343 carbapenemase-positive isolates within these non-human categories. These patterns reflect deposited projects and uneven sectoral sampling rather than sector-specific AMR prevalence (Table 4; Fig 4).

**Fig 4.**
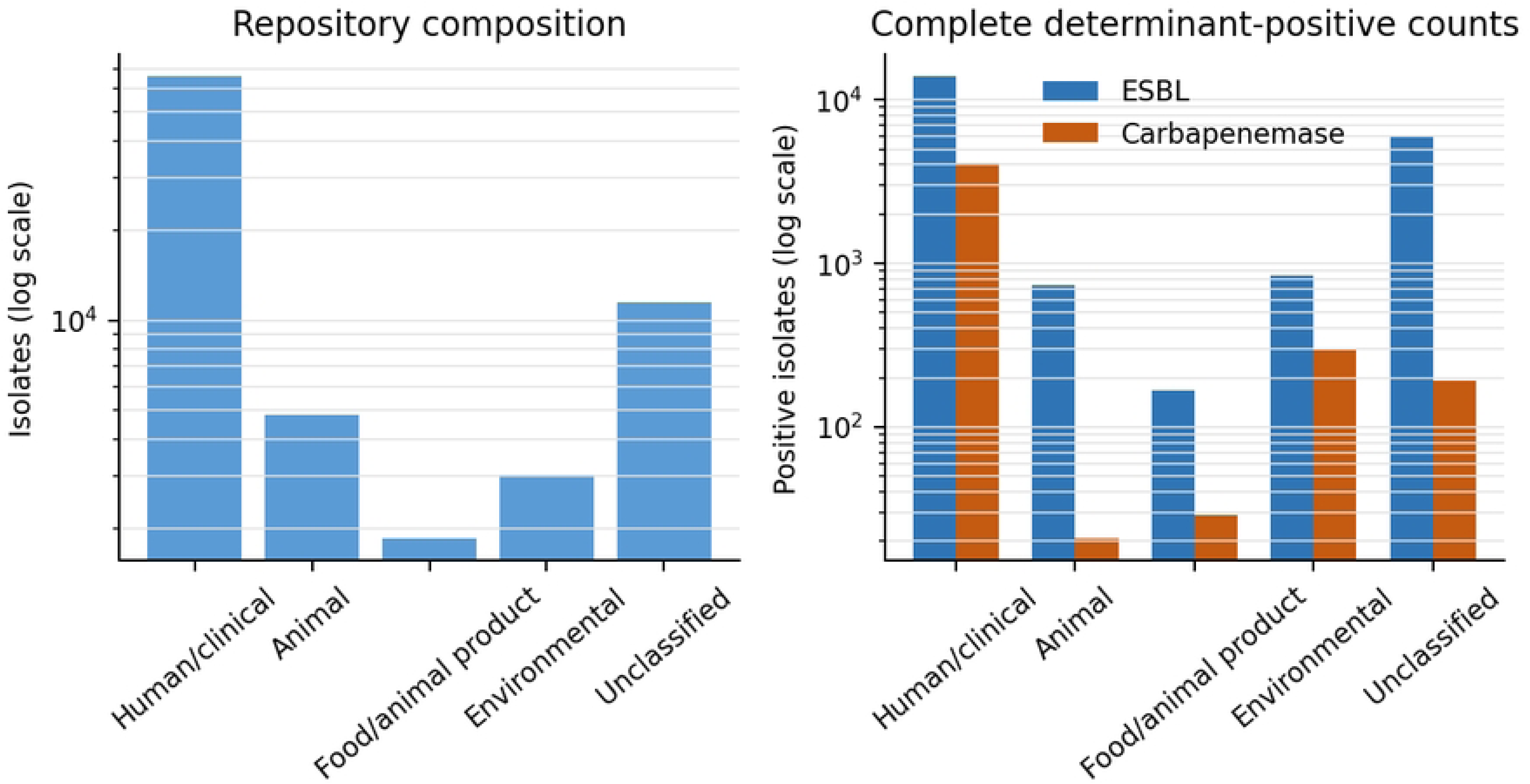
One Health source composition and complete determinant-positive counts.

**Table 4.** Repository composition and complete AMR outcomes by harmonised One Health source category.

| Source category | Isolates | Africa share | ESBL + | Carbapenemase + | mcr + | mecA + |
| --- | --- | --- | --- | --- | --- | --- |
| Human/clinical | 65,665 | 75.6% | 13,819 (21.0%) | 4,107 (6.3%) | 157 | 833 |
| Animal | 4,803 | 5.5% | 735 (15.3%) | 21 (0.4%) | 54 | 32 |
| Food/animal product | 1,864 | 2.2% | 168 (9.0%) | 29 (1.6%) | 14 | 26 |
| Environmental | 3,024 | 3.5% | 845 (27.9%) | 293 (9.7%) | 16 | 7 |
| Unclassified | 11,473 | 13.2% | 5,946 (51.8%) | 192 (1.7%) | 21 | 37 |

*Note: Source categories were inferred from multiple metadata fields and are analytical harmonisations rather than official NCBI categories.*

### 3.4 Subregional and regional-economic-community patterns

Repository size did not correspond to determinant frequency. Eastern Africa contributed 40.6% of isolates, whereas Northern Africa contributed only 4.8% but had the highest carbapenemase frequency (32.3%). The Northern African signal was concentrated in selected Egyptian and Tunisian projects and organism groups (Fig 5). In the REC analysis, SADC contained the largest collection, while UMA and CEN-SAD had the highest carbapenemase frequencies. Because REC memberships overlap, these totals are non-additive and represent policy groupings rather than independent regional estimates (Table 5).

**Fig 5.**
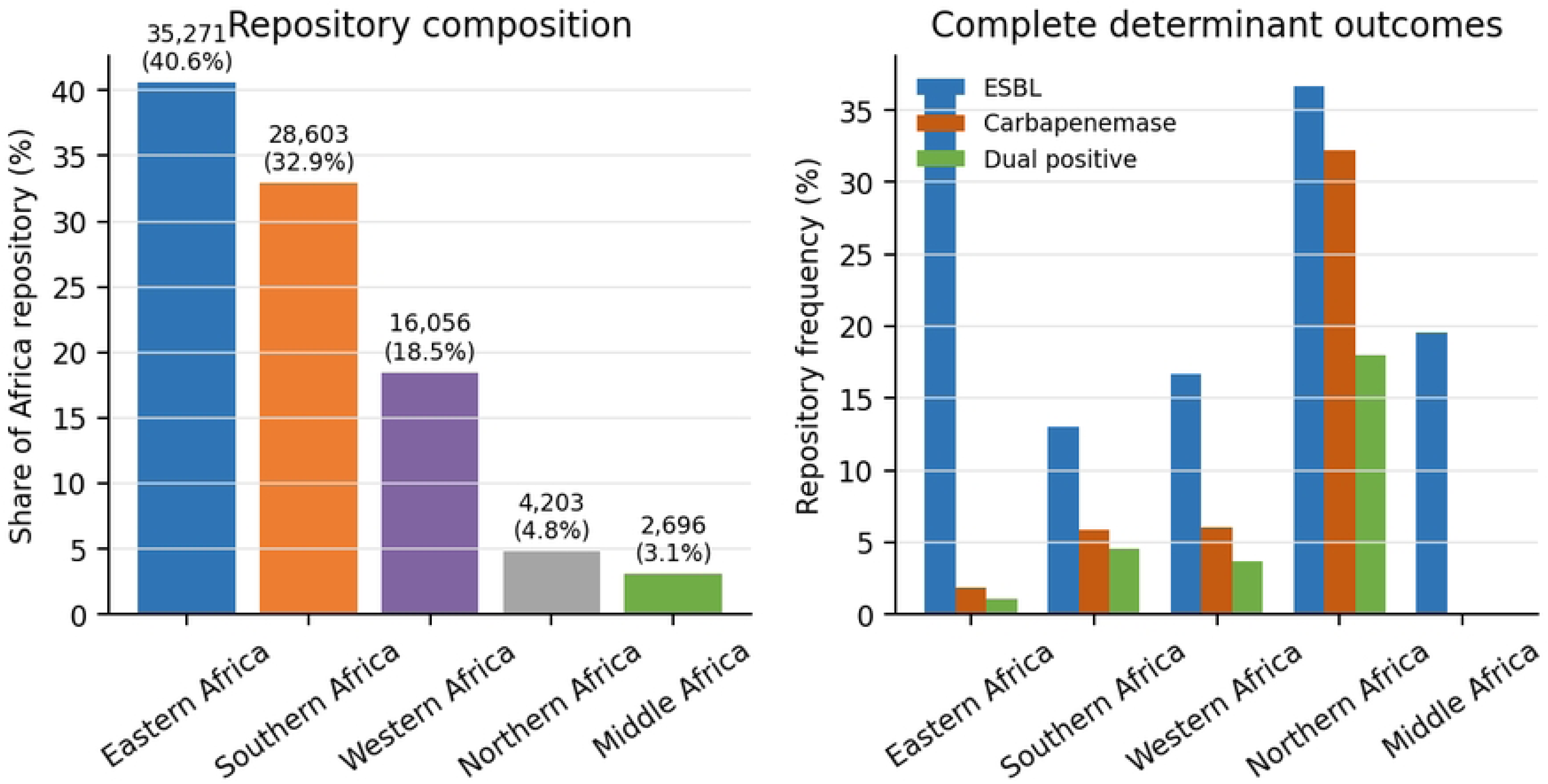
Repository composition and complete determinant outcomes by UN M49 subregion.

**Table 5.** Secondary analysis by African Union-recognised regional economic community.

| Regional economic community | Isolates | Africa share | Countries | ESBL + | Carbapenemase + |
| --- | --- | --- | --- | --- | --- |
| SADC | 50,888 | 58.6% | 14 | 11,578 (22.8%) | 1,794 (3.5%) |
| COMESA | 34,880 | 40.2% | 20 | 13,214 (37.9%) | 1,827 (5.2%) |
| CEN-SAD | 20,097 | 23.2% | 25 | 4,317 (21.5%) | 2,284 (11.4%) |
| EAC | 17,071 | 19.7% | 8 | 5,332 (31.2%) | 501 (2.9%) |
| IGAD | 14,240 | 16.4% | 8 | 5,097 (35.8%) | 566 (4.0%) |
| ECOWAS | 11,749 | 13.5% | 12 | 2,253 (19.2%) | 733 (6.2%) |
| ECCAS | 3,092 | 3.6% | 10 | 756 (24.5%) | 15 (0.5%) |
| UMA | 2,366 | 2.7% | 5 | 842 (35.6%) | 550 (23.3%) |

*Note: REC memberships overlap. A country contributes to every REC of which it is a current member, so rows are not additive or statistically independent.*

### 3.5 Multicountry genomic-cluster signals

Among 675 multicountry SNP-cluster roots, 335 spanned more than one UN M49 subregion. Of the 118 clusters containing a complete carbapenemase, 70 included carbapenemase-positive isolates from at least two countries. The largest was *A. baumannii* cluster PDS000005671, comprising 202 carbapenemase-positive isolates from eight countries across three subregions (Table 6).

**Table 6.** Leading carbapenemase-bearing multicountry SNP-cluster candidates.

| Cluster root | Organism | Africa n | Countries | Subregions | Carb + | Complete carbapenemases |
| --- | --- | --- | --- | --- | --- | --- |
| PDS000005671 | *Acinetobacter baumannii* | 202 | 8 | 3 | 202 | *blaNDM*; *blaNDM-1*; *blaNDM-4*; *blaOXA-23* |
| PDS000054886 | *Pseudomonas aeruginosa* | 112 | 8 | 3 | 98 | *blaNDM-1*; *blaVIM-2*; *blaVIM-5* |
| PDS000051203 | *Klebsiella pneumoniae* | 75 | 2 | 1 | 69 | *blaKPC-2*; *blaNDM-1*; *blaNDM-5*; *blaOXA-181*; *blaOXA-48* |
| PDS000085592 | *Klebsiella pneumoniae* | 52 | 2 | 1 | 50 | *blaNDM*; *blaNDM-5*; *blaOXA-48* |
| PDS000265951 | *Acinetobacter baumannii* | 46 | 6 | 2 | 45 | *blaNDM-1*; *blaOXA-23* |
| PDS000268214 | *E. coli and Shigella* | 188 | 11 | 3 | 33 | *blaNDM-1*; *blaNDM-5*; *blaOXA-181* |
| PDS000045135 | *Klebsiella pneumoniae* | 34 | 2 | 1 | 33 | *blaNDM*; *blaNDM-1*; *blaOXA-48* |
| PDS000019298 | *E. coli and Shigella* | 30 | 3 | 2 | 30 | *blaNDM-1*; *blaNDM-5*; *blaOXA-181* |
| PDS000264503 | *Acinetobacter baumannii* | 28 | 4 | 1 | 28 | *blaNDM*; *blaNDM-1*; *blaNDM-9*; *blaOXA-23* |
| PDS000006578 | *Klebsiella pneumoniae* | 28 | 2 | 2 | 28 | *blaNDM-1*; *blaOXA-181*; *blaOXA-48* |

Note: Clusters are ranked primarily by carbapenemase-positive isolate count. Cluster membership is an NCBI relatedness signal; official trees, pairwise distances, dates, sources, and epidemiological links are required for transmission assessment.

Existing REC memberships covered all countries involved in only 51.7% of the broader clusters and 54.3% of the strict carbapenemase-positive clusters. Although 68.3% of strict country-pair connections involved countries sharing an REC, sampling-normalised overlap remained low, reaching a maximum Jaccard index of 9.5% for Libya–Tunisia (Table 7; Fig 6). These clusters identify priorities for tree-based and epidemiological investigation but do not establish transmission.

**Fig 6.**
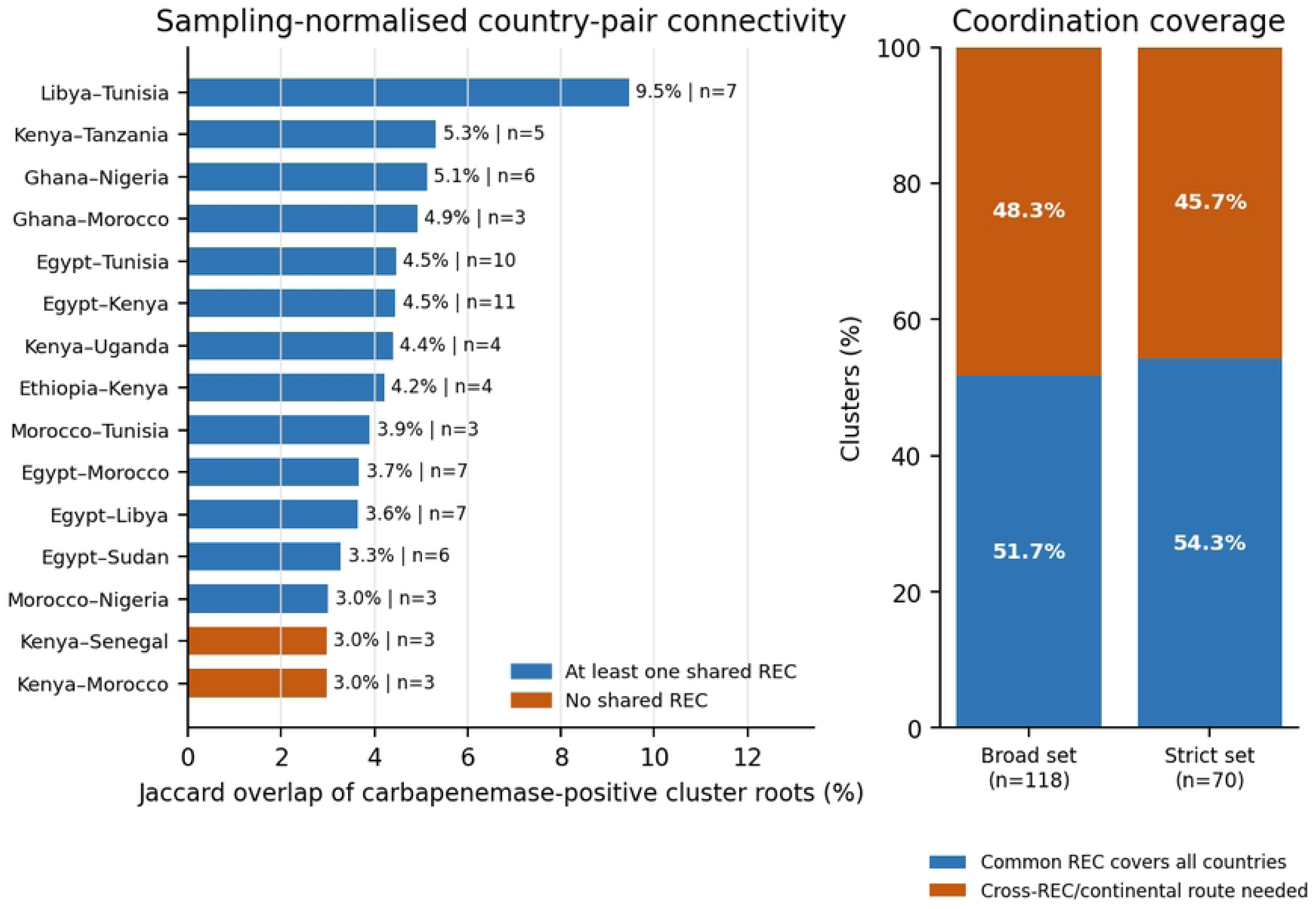
Sampling-normalised carbapenemase-cluster connectivity and REC coordination coverage.

**Table 7.** Leading country-pair genomic-connectivity signals.

| Country pair | Shared carb + clusters | Carb-cluster Jaccard | All shared clusters | Shared REC(s) | Subregions |
| --- | --- | --- | --- | --- | --- |
| Egypt–Kenya | 11 | 4.5% | 27 | COMESA | Different |
| Egypt–Tunisia | 10 | 4.5% | 18 | CEN-SAD; COMESA | Same |
| Libya–Tunisia | 7 | 9.5% | 8 | CEN-SAD; COMESA; UMA | Same |
| Egypt–Morocco | 7 | 3.7% | 11 | CEN-SAD | Same |
| Egypt–Libya | 7 | 3.6% | 8 | CEN-SAD; COMESA | Same |
| Ghana–Nigeria | 6 | 5.1% | 35 | CEN-SAD; ECOWAS | Same |
| Egypt–Sudan | 6 | 3.3% | 7 | CEN-SAD; COMESA | Same |
| Kenya–Tanzania | 5 | 5.3% | 100 | EAC | Same |
| Egypt–Nigeria | 5 | 2.0% | 11 | CEN-SAD | Different |
| Kenya–Uganda | 4 | 4.4% | 99 | COMESA; EAC; IGAD | Same |
| Ethiopia–Kenya | 4 | 4.2% | 37 | COMESA; IGAD | Same |
| Egypt–South Africa | 4 | 1.3% | 13 | None | Different |

*Note: A strict carbapenemase-positive connection required both countries to contribute at least one carbapenemase-positive isolate to the same cluster. Jaccard values divide shared roots by the union of eligible roots in the two countries. Connectivity is not evidence of direct transmission.*

### 3.6 BioProject sensitivity and metadata completeness

The 86,829 isolates represented 2,736 BioProjects, but project concentration varied substantially between subregions. Southern Africa was the most concentrated, with its largest project contributing 39.6% of isolates. Project balancing reduced the Eastern African ESBL estimate from 36.9% to 12.5% and the Northern African carbapenemase estimate from 32.3% to 17.9%. Changes were not uniform across subregions, demonstrating the influence of project and sample composition on raw repository frequencies (Table 8; Fig 7).

**Fig 7.**
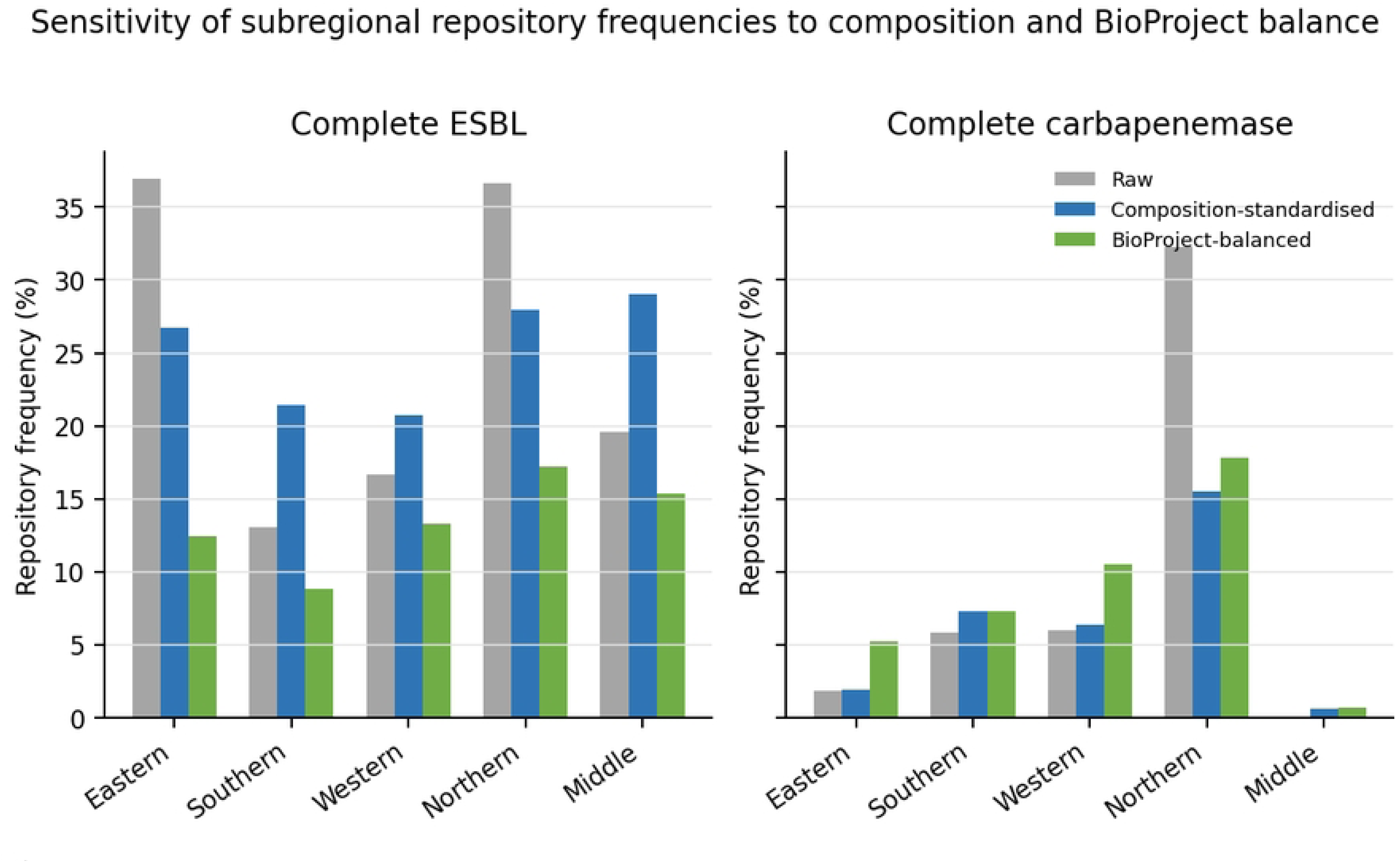
Sensitivity of subregional repository frequencies to sample composition and BioProject balance.

**Table 8.** BioProject concentration and standardised sensitivity estimates by subregion.

| UN M49 subregion | Largest project share | Effective projects | Raw ESBL | Project-balanced ESBL | Raw carb | Project-balanced carb |
| --- | --- | --- | --- | --- | --- | --- |
| Eastern Africa | 11.6% | 31.32 | 36.9% | 12.5% | 1.8% | 5.3% |
| Southern Africa | 39.6% | 5.33 | 13.1% | 8.9% | 5.8% | 7.3% |
| Western Africa | 10.8% | 44.42 | 16.7% | 13.3% | 6.0% | 10.6% |
| Northern Africa | 6.7% | 58.2 | 36.7% | 17.2% | 32.3% | 17.9% |
| Middle Africa | 13.1% | 15.79 | 19.6% | 15.4% | 0.1% | 0.7% |

*Note: Effective projects equal the inverse BioProject Herfindahl-Hirschman index. Project-balanced values derive from regularised models that give each BioProject equal total weight and standardise organism, source and collection period. They are descriptive sensitivity estimates, not corrected prevalence.*

Metadata completeness was also uneven. Collection date and isolation source were available for 89.3% and 78.4% of isolates, respectively, whereas geographic coordinates were available for 27.5% and structured source_type for only 5.0%. Completeness also varied considerably between countries (Fig 8).

**Fig 8.**
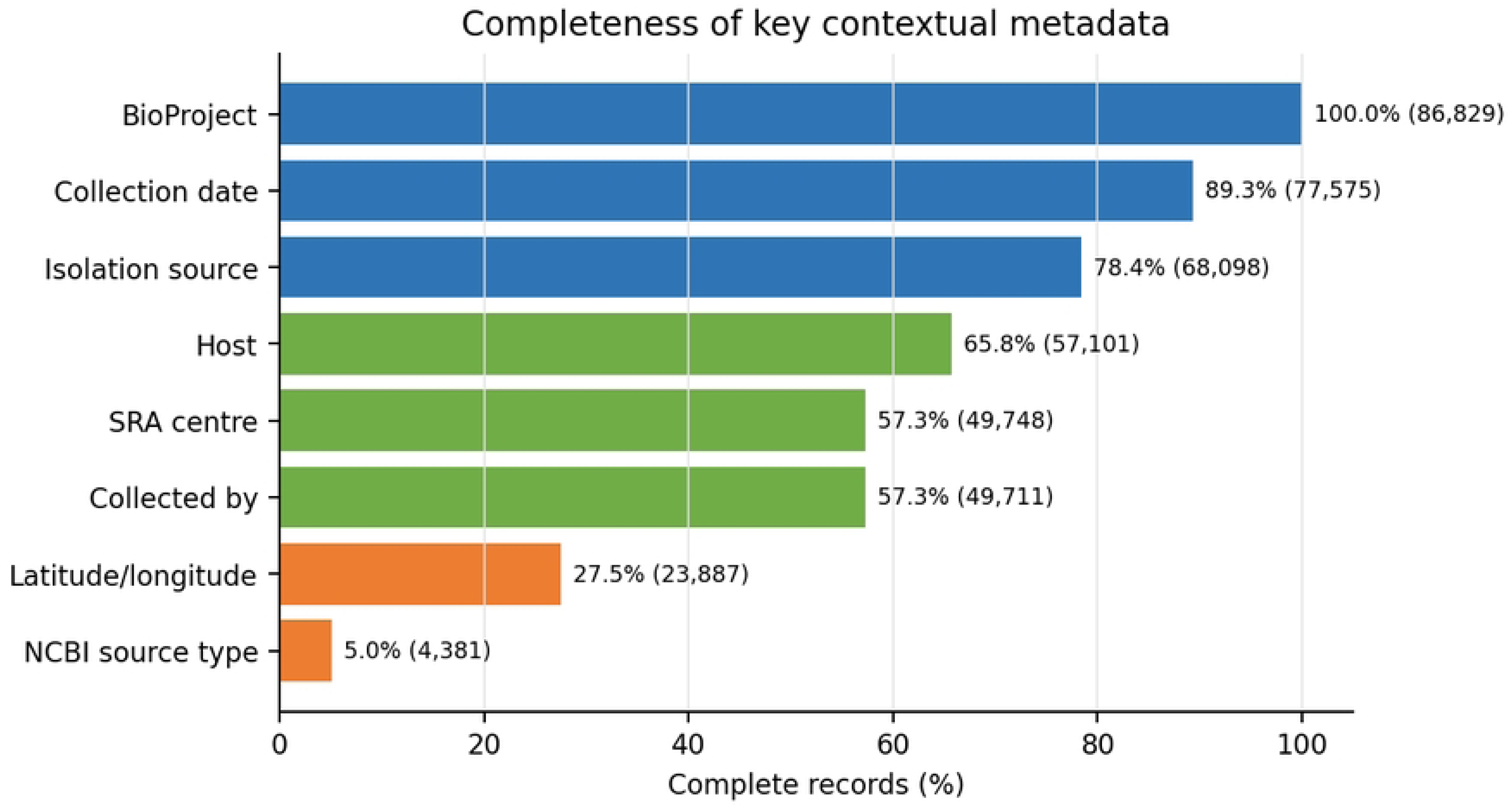
Overall completeness of key contextual metadata fields.

### 3.7 Regional policy mechanisms

Documented coordination mechanisms differed across RECs. The EAC and SADC had AMR-specific regional mechanisms; ECOWAS and IGAD incorporated AMR within laboratory, One Health or regulatory programmes; and COMESA and ECCAS had broader health and laboratory structures. No REC-specific AMR or genomic-surveillance mechanism was identified for CEN- SAD or UMA, although both contained strict carbapenemase-sharing clusters (Table 9).

**Table 9.** Genomic-cluster signals and documented coordination mechanisms in the eight AU- recognised RECs.

| REC | Members represented | Shared SNP clusters | Strict carb + clusters | Documented mechanism category | Documented entry point |
| --- | --- | --- | --- | --- | --- |
| COMESA | 20 | 294 | 44 | Regional health/laboratory mechanism; no REC-wide AMR framework identified | COMESA Health Desk, reference/satellite laboratories and SPS technical working groups |
| SADC | 14 | 232 | 4 | AMR-specific regional framework | SADC health programme and Member State AMR structures |
| EAC | 8 | 207 | 8 | AMR-specific regional laboratory programme | EAC Regional Network of Public Health Reference Laboratories and LIMS |
| CEN-SAD | 25 | 171 | 37 | No REC-specific mechanism identified; this is not evidence of absence | Africa CDC AMRSNET/RISLNET and overlapping RECs |
| IGAD | 8 | 134 | 8 | One Health/health-regulatory mechanism explicitly addressing AMR | IGAD One Health coordination and Medicines Regulatory Harmonization expert groups |
| ECOWAS | 12 | 71 | 7 | Regional health/laboratory network with explicit AMR component | West African Health Organisation, PROALAB and regional laboratory data platforms |
| UMA | 5 | 23 | 12 | No REC-specific mechanism identified; this is not evidence of absence | Africa CDC AMRSNET/RISLNET, WHO regional structures and overlapping COMESA/CEN-SAD mechanisms |
| ECCAS | 10 | 15 | 0 | Regional health/laboratory and One Health mechanism; no REC-wide AMR framework identified | OSAC surveillance and One Health departments and Central Africa RISLNET |

*Note: REC memberships overlap and counts are non-additive. Strict clusters contain carbapenemase-positive isolates from at least two member states. Documented mechanisms do not indicate implementation or effectiveness. Sources: [20,24-30].*

## 4 Discussion

This analysis showed that Africa’s public genomic repository contained important but unevenly represented AMR signals. *bla*CTX-M-15 dominated the ESBL profile, while NDM and OXA enzymes were the principal carbapenemases. These determinants were concentrated in *K. pneumoniae*, *A. baumannii* and *P. aeruginosa*, and nearly two-thirds of carbapenemase-positive isolates also carried an ESBL. Multicountry clusters were common, but low sampling-normalised overlap and substantial sensitivity to BioProject composition showed that these findings should be interpreted as surveillance priorities rather than national or continental prevalence estimates. The predominance of *bla*CTX-M-15 is consistent with studies showing its circulation across genetically diverse African *E. coli* populations and clinical collections [31,32]. The NDM and OXA profiles also agree with reports from Chad, Egypt, North Africa and South Africa [33–37]. Their concentration in *K. pneumoniae* and non-fermenting Gram-negative organisms is clinically important because carbapenem-resistant Enterobacterales and *A. baumannii* are WHO critical-priority pathogens. In contrast, carbapenem-resistant *P. aeruginosa* remains a high priority [38]. The high ESBL–carbapenemase co-detection indicates the accumulation of multiple β-lactam-resistance mechanisms in the same deposited isolates and may further restrict treatment options. Such isolates warrant phenotypic susceptibility and plasmid-context analysis because genomic detection alone does not establish gene expression, transmissibility or treatment failure. Detection of similar determinants in hospital wastewater also supports surveillance beyond clinical settings [39].

Organism and source patterns further refined surveillance priorities. *K. pneumoniae* combined a high ESBL repository frequency (80.6%) with 19.7% ESBL–carbapenemase co-detection, while carbapenemases were frequent in *A. baumannii* (73.9%) and *P. aeruginosa* (30.9%); almost all *mecA* detections occurred in *S. aureus* (Table 3). These patterns support organism-specific genomic and phenotypic follow-up but, because they reflect the projects and organisms deposited, they are not species-level prevalence estimates. Source representation was similarly uneven: human or clinical isolates dominated, while animal, food and environmental sources together accounted for only 11.1%, and 293 of the 343 carbapenemase-positive isolates in these non-human categories came from environmental samples concentrated in targeted projects (Table 4). This cannot be interpreted as higher environmental prevalence; rather, it demonstrates that current repository coverage is inadequate for sectoral comparison and supports more representative One Health sampling [7,9].

The multicountry clusters provide a shortlist for further investigation, not evidence of cross-border transmission. Although 70 clusters contained carbapenemase-positive isolates from at least two countries, country-pair Jaccard overlap remained low after accounting for each country’s cluster volume. NCBI cluster membership indicates genomic relatedness but does not establish the timing, direction or setting of transmission [12]. Official trees, pairwise SNP distances, collection dates, sources and epidemiological information are therefore required to determine whether these signals represent recent clonal spread, older shared ancestry or unrelated acquisition of similar resistance determinants.

Mapping the clusters to RECs identified possible routes for regional follow-up. In 38 of the 70 strict clusters, all participating countries shared at least one REC; the remaining 32 crossed REC boundaries and would require cross-REC or continent-wide coordination. Because several large clusters spanned multiple subregions and RECs, continental coordination is needed to assemble the country-specific trees, dates, source data and phenotypes required for investigation; this does not imply that the clusters represent transcontinental transmission. The policy review identified AMR-specific mechanisms in the EAC and SADC and laboratory, One Health or health- coordination structures in ECOWAS, IGAD, COMESA and ECCAS through which cluster investigations could be organised [24–28,30]. No REC-specific mechanism was identified for CEN-SAD or UMA in the official documents reviewed [20,29]. This reflects the official-source search and does not establish that no relevant activities exist. This documentary gap supports the use of Africa CDC’s AMRSNET and RISLNET as continental pathways for signals not covered by a common REC [40,41].

Repository representation was highly unequal. Three countries contributed more than half of all isolates, while several states had few or no qualifying records. Subregional frequencies also changed substantially after organism, source and BioProject composition were considered. Project balancing reduced the Eastern African ESBL estimate from 36.9% to 12.5% and the Northern African carbapenemase estimate from 32.3% to 17.9%. These shifts show that raw repository frequencies can be driven by a small number of targeted projects and should not be used to rank subregional AMR burden. Similar inequalities in sequencing, bioinformatics and data production have been reported elsewhere in Africa [14,15,17]. Collaborative initiatives such as SeqAfrica can strengthen locally usable genomic surveillance, but sustained national capacity is needed to reduce dependence on episodic or externally led sequencing projects [16].

Incomplete metadata further limited interpretation. Although collection date and isolation source were available for 89.3% and 78.4% of records, geographic coordinates were available for only 27.5% and structured source-type metadata for 5.0%. One Health categories therefore often had to be inferred from free text, introducing possible misclassification. Standardised contextual metadata are necessary to distinguish human, animal, food and environmental pathways and interpret genomic relatedness [9]. Frameworks such as PHA4GE provide practical standards for improving repository submissions and data reuse [42]. More broadly, repository isolates are not probability samples, and the sensitivity models expose project-related instability rather than correct it. Genotypic calls were not systematically linked to phenotypic susceptibility, clinical outcomes or plasmid context, while cluster assignments may change as NCBI updates its pipeline. The policy analysis assessed published mechanisms rather than their implementation. WGS findings should therefore complement phenotypic and epidemiological surveillance [7].

The findings support coordinated surveillance at national, REC and continental levels. National programmes should prioritise organism-specific monitoring of *bla*CTX-M-15, NDM-family enzymes and clinically important OXA carbapenemases with phenotypic confirmation and epidemiological review. Multicountry clusters within a common REC can undergo joint tree analysis and metadata review through existing regional mechanisms, while cross-REC signals should be coordinated through Africa CDC. Across all levels, investment should prioritise underrepresented countries, non-human sources, standardised metadata and locally sustained sequencing and bioinformatics capacity.

## 5 Conclusion

Public repositories identified widespread *bla*CTX-M-15, NDM and OXA resistance signals across Africa, but uneven sampling and BioProject composition preclude prevalence interpretation. Seventy carbapenemase-positive multicountry clusters were detected; their low normalised overlap indicated genomic relatedness rather than recent transmission. Existing RECs could coordinate 38 clusters, while 32 required cross-REC or Africa CDC coordination. Effective surveillance therefore requires national phenotypic and epidemiological confirmation, regional cluster review, representative One Health sampling, standardised metadata and sustained African sequencing and bioinformatics capacity.

## Data availability statement

The genomic sequences and contextual metadata analysed in this study are publicly available through NCBI Pathogen Detection, BioSample and the Sequence Read Archive. The analysis snapshot was retrieved on 24 August 2026. The accompanying reproducibility package is organised as S1–S5 Files and contains the raw repository snapshot, the cleaned and deduplicated analysis-ready isolate dataset, analytical result tables and policy-source matrix, reproducibility code, and the geographic-validation, BioSample-deduplication and provenance audit. BioSample, assembly, BioProject and SNP-cluster identifiers are retained to link analytical records to the source repositories.

## Ethics statement

This study used publicly accessible, non-identifiable bacterial isolate records and did not involve participant recruitment, intervention or access to individual clinical records. Institutional ethical approval was therefore not required.

## Author contributions

RN and GNM conceptualised the study and developed the methodology. RN was responsible for data curation, formal analysis, and drafting the original manuscript. GNM contributed to the formal analysis and participated in the critical review and editing of the manuscript. FOF contributed to the review and editing of the final manuscript. All authors have read and agreed to the published version of the manuscript.

## Declaration of generative AI use

During preparation of this work, the authors used ChatGPT (OpenAI) and Google AI to assist with Python code development and manuscript organisation, and Grammarly for language editing. All code, outputs, interpretations and text were reviewed and verified by the authors, who take full responsibility for the work.

## Data Availability

The genomic sequences and contextual metadata analysed in this study are publicly available through NCBI Pathogen Detection, BioSample and the Sequence Read Archive. The analysis snapshot was retrieved on 24 August 2026. The accompanying reproducibility package is organised as S1–S5 Files and contains the raw repository snapshot, cleaned and analysis-ready isolate-level dataset, analytical result tables and policy-source matrix, reproducibility code, and geographic-validation, BioSample-deduplication and provenance audit. BioSample, assembly, BioProject and SNP-cluster identifiers are retained to link analytical records to the source repositories.

## Supporting information

S1 File. Raw NCBI Pathogen Detection snapshot.

S2 File. Cleaned, deduplicated and analysis-ready isolate-level dataset.

S3 File. Analytical result tables and policy-source matrix.

S4 File. Reproducibility code for the Pan-African genomic AMR repository analysis. S5 File. Geographic validation, BioSample deduplication and provenance audit.

